# The prevalence and density of asymptomatic *Plasmodium falciparum* infections among children and adults in three communities of western Kenya

**DOI:** 10.1101/2021.03.31.21254671

**Authors:** Christina Salgado, George Ayodo, Michael D. Macklin, Meetha P. Gould, Srinivas Nallandhighal, Eliud O. Odhiambo, Andrew Obala, Wendy O’Meara Prudhomme, Chandy C. John, Tuan M. Tran

## Abstract

**Background:** Further reductions in malaria incidence as more countries approach malaria elimination require the identification and treatment of asymptomatic individuals who carry mosquito-infective *Plasmodium* gametocytes that are responsible for furthering malaria transmission. Assessing the relationship between total parasitemia and gametocytemia in field surveys can provide insight as to whether detection of low-density, asymptomatic *Plasmodium falciparum* infections using sensitive molecular methods can sufficiently detect the majority of infected individuals who are potentially capable of onward transmission.

**Methods:** In a cross-sectional survey of 1,354 healthy children and adults in three communities in western Kenya across a gradient of malaria transmission (Ajigo, Webuye, and Kapsisywa-Kipsamoite), we screened for asymptomatic *P. falciparum* infections by rapid diagnostic tests, blood smear, and quantitative PCR of dried blood spots targeting the *varATS* gene in genomic DNA. A multiplex quantitative reverse-transcriptase PCR assay targeting female and male gametocyte genes (*pfs25, pfs230p*), a gene with a transcriptional pattern restricted to asexual blood-stages (*piesp2*), and human *GAPDH* was also developed to determine total parasite and gametocyte densities among parasitemic individuals.

**Results:** The prevalence of *varATS*-detectable asymptomatic infections was greatest in Ajigo (42%), followed by Webuye (10%). Only two infections were detected in Kapsisywa. No infections were detected in Kipsamoite. Across all communities, children aged 11-15 years account for the greatest proportion total and sub-microscopic asymptomatic infections. In younger age groups, the majority of infections were detectable by microscopy, while 68% of asymptomatically infected adults (>21 years old) had sub-microscopic parasitemia. *Piesp2*-derived parasite densities correlated poorly with microscopy-determined parasite densities in patent infections relative to *varATS*-based detection. In general, both male and female gametocytemia increased with increasing *varATS*-derived total parasitemia. A substantial proportion (41.7%) of individuals with potential for onward transmission had qPCR-estimated parasite densities below the limit of microscopic detection but above the detectable limit of *varATS* qPCR.

**Conclusions:** This assessment of parasitemia and gametocytemia in three communities with different transmission intensities revealed evidence of a substantial sub-patent infectious reservoir among asymptomatic carriers of *P. falciparum*. Experimental studies are needed to definitively determine whether the low-density infections in communities such as Ajigo and Webuye contribute significantly to malaria transmission.

## INTRODUCTION

Malaria remains a global public health burden with more than 200 million cases worldwide [1]. Transmission of malaria requires that sexual-stage *Plasmodium* parasites, gametocytes, present in the blood of infected humans be ingested by female *Anopheles* mosquitoes during feeding. Strategies that combine effective control of the mosquito vector through use of insecticide-treated nets (ITNs) and indoor residual spraying alongside rapid diagnosis and effective treatment of malaria with artemisinin-combination therapy (ACT) have reduced the prevalence of *Plasmodium falciparum* infection and the incidence of clinical malaria in endemic areas of Africa since 2000, albeit at a slower rate in recent years [1]. Further reductions in malaria incidence as more countries approach malaria elimination would require the identification and treatment of asymptomatic individuals who carry mosquito-infective gametocytes that are responsible for furthering malaria transmission [2].

Detection of asymptomatically infected individuals has been a major challenge given that individuals residing in areas of high-transmission intensity often carry parasitemia at densities below the detection limits of accessible field diagnostics, which currently includes microscopy and rapid-diagnostic tests (RDTs) [3]. Moreover, the proportion of low-density infections among all malaria infections in a community increases with decreasing malaria transmission [2], suggesting that more sensitive diagnostics are required for detecting parasitemia among individuals in low-transmission settings [4]. Several studies have examined whether low-density infections contribute to onward transmission using mosquito feeding assays [5-11]. A recent meta-analysis of eight such studies estimated that individuals with sub-patent parasitemia were approximately one-third as infectious to mosquitoes as individuals with blood-smear positive infections [4]. In general, gametocyte density directly correlates with mosquito infectivity and thus transmission, with infections with parasite densities below the limit of detection of conventional molecular diagnostics being unlikely to contribute significantly to transmission [12]. Assessing the relationship between total parasitemia and gametocytemia in field surveys can provide insight as to whether detection of low-density, asymptomatic *P. falciparum* infections using sensitive molecular methods can sufficiently identify the majority of infected individuals who are potentially capable of onward transmission.

In this study, we determined the prevalence and density of asymptomatic *P. falciparum* infections among children and adults in three communities of western Kenya that differed in transmission intensities using quantitative molecular assays. To better estimate the relationship between asexual parasite densities and gametocyte densities, we sought to develop a multiplex quantitative reverse-transcriptase PCR assay that could detect asexual-stage specific, female gametocyte-specific, and male gametocyte-specific genes in a single blood sample. Results were compared to microscopy and an established quantitative PCR-based diagnostic assay.

## METHODS

### Ethics Statement

The study was reviewed and approved by the Kenya Medical Research Institute Ethics Review Committee and the Indiana University Institutional Review Board. Written informed consent was obtained from a parent or guardian of participants who were minors and from adult participants. Minors ≥ 15 years of age provided their own written informed assent, accompanied by written consent of a parent or guardian.

### Study Sites and Study Participants

The study was conducted from August to September 2016 at three sites in western Kenya that differed in malaria transmission intensity. Ajigo, located in the lowland area of Siaya County, where malaria transmission is typically intense [13]. Webuye Town, a town located in Bungoma County, exhibits moderate, perennial transmission with seasonal peaks from May to June, and the study area has been previously described [14-16]. Kipsamoite and Kapsisywa, two adjacent highland communities in Nandi County, have low and unstable malaria transmission that has recently been described in detail [17, 18]. Healthy participants aged 1 to 85 years were sampled from a randomized community census of households for each site and enrolled over a four-week period. A brief questionnaire that included gender, age, recent travel history within the last month, recent use of ITNs, and relevant medical history was administered. Exclusion criteria at enrollment were axillary temperature ≥ 37.5°C, known acute systemic or chronic illness, use of antimalarial or immunosuppressive medications within 30 days prior, or pregnant at time of survey.

### Blood Collection

Drops of blood were collected by fingerprick for RDT (Paracheck Pf; Orchid Biomedical Systems), whole-blood RNA, thick and thin blood smears, and dried blood spots (DBS) on filter paper (903 Protein Saver; Whatman). Individuals who tested positive for asymptomatic *P. falciparum* infection by RDT were treated at the point-of-care using the standard regimen recommended by the Ministry of Health in Kenya. For whole-blood RNA, 200 μl of peripheral fingerprick blood was collected using capillary blood collection tubes containing EDTA (Microvette CB300 K2E; Sarstedt) and transferred immediately in cryotubes pre-filled with 400 μl Tempus solution (Applied Biosystems). Filled sample tubes were agitated vigorously per the manufacturer’s instructions and stored at -80°C within 24 hours of collection until use.

### Microscopy

Giemsa-stained blood smears were examined for the presence of asexual parasites in 200 fields using the 100x oil immersion objective lens by two trained microscopists. Independent verification was performed by a third reader for samples that were discordant between the first two microscopists. For positive samples, the number of asexual parasites per 200 leukocytes was multiplied by 40 to convert to parasites per μl, assuming an average leukocyte count of 8,000 leukocytes per μl of blood.

### Parasite Culture

To produce parasite gDNA for use in standard curves for parasite density determination, *P. falciparum* 3D7 parasites (Malaria Research and Reference Reagent Resource Center [MR4], BEI Resources) were cultured *in vitro* using standard techniques [19] with two rounds of synchronization by sorbitol treatment to achieve a high parasitemia. Ring and early trophozoite stage *P. falciparum* parasites were 10-fold serially diluted in whole blood of an uninfected North American donor to obtain a final density of 440,000 down to 0.44 parasites/µl and spotted on 903 Protein Saver cards. To produce parasite RNA for use in standard curves for gametocyte density estimates, *P. falciparum* NF54 (MR4) *in vitro* cultures were enriched for gametocytes by decreasing asexual parasitemia [16]. Total gametocytes (without differentiating for sex) were counted and 10-fold serially diluted in whole blood to obtain a final density of 2580 down to 0.00258 gametocytes/μl immediately prior to RNA stabilization with Tempus solution at a 1:2 ratio. Asexual parasites (rings, trophozoites, and schizonts) were also counted in the same culture to allow parasite quantification using asexual-stage specific targets.

### DNA and RNA isolation

Total DNA was extracted from three 0.32 cm diameter circles punched from each DBS using the QIAamp 96 Blood Kit (Qiagen, Valencia, CA) per the manufacturer’s instructions and eluted in 50 µl Tris-EDTA buffer. RNA was extracted from whole-blood RNA in Tempus using Norgen RNA extraction kit (Norgen Biotek, Thorold, Ontario) and treated with RNase-Free DNase I Kit (cat 25710) to a final elution volume of 50 µl, per manufacturer’s instructions. Extracted RNA samples were assessed for quality and quantity using automated parallel capillary electrophoresis (Fragment Analyzer System, Agilent).

### Real-time quantitative PCR using genomic DNA

To detect the presence of *P. falciparum* genomic DNA isolated from the DBS, we adapted primers targeting *P. falciparum varATS* that were originally designed for use in a Taqman-based qPCR assay [20] for use with the PowerUp SYBR Green Master Mix System (Thermo Fisher Scientific, Waltham, Massachusetts) (**Table S1**). Samples were assayed in triplicate in 384-well plates on a QuantStudio 6 Flex Real Time PCR System (Thermo Fisher Scientific, Waltham, Massachusetts) using standard cycling conditions and a melt curve analysis. During assay development, we verified that wells with a first calculated melt temperature (Tm1) >71.14°C contained *varATS* amplicons by Sanger sequencing, whereas wells with Tm1 <71.14°C contained primer dimers. Subsequently, we set the criteria for a *P. falciparum* positive sample as having ≥2 of 3 replicate wells with a Ct <39 AND a Tm1 >71.14°C. Using these criteria, genomic DNA samples isolated from the blood of 20 of 20 (100%) healthy North American controls with no malaria exposure history were confirmed to be *P. falciparum* negative, and 86 of 87 (98.9%) samples positive by conventional *P. falciparum* 18s rRNA PCR [21, 22] were confirmed as positive with the modified *varATS*-based assay. The one discordant sample had only 1 of 3 replicate wells meeting the Ct and Tm1 criteria. A standard curve of gDNA extracted from serially diluted *P. falciparum*-spiked DBS samples (described above) and no-template negative controls were run on every plate, which allowed for estimation of parasite densities using Ct values.

### Multiplex real-time quantitative reverse transcription PCR

We initially sought to develop a four-plex real-time quantitative reverse transcription PCR (RT-qPCR) that would detect female gametocytes, male gametocytes, and asexual parasites, as well as a human housekeeping gene glyceraldehyde 3-phosphate dehydrogenase (GAPDH), which served as a control for RNA extraction and relative quantification. The genes *pfs25* [23] and *pfs230p* were used as the female- and male-specific gametocyte targets, respectively. The gene encoding for parasite-infected erythrocyte surface protein (*piesp2*, also called PFE60 and PF3D7_0501200) was chosen based on a transcriptional pattern restricted to asexual blood-stages, particularly trophozoites, in three *P. falciparum* gene expression datasets available on PlasmoDB (http://plasmodb.org) [24-26]. Primers and probes for *pfs25* were adapted from Wampfler et al [27]. Primers and probes for *pfs230p* and *piesp2* were developed *de novo* using Primer 3 software [28] following standard guidelines for qPCR primer design. All primer and probes are listed in **Table S1**.

After generation of cDNA from 50 ng RNA for each sample replicate using LunaScript RT Supermix (New England Biolabs) under standard cycling conditions, 20X triplex master mix was prepared from appropriate final concentrations of primers and probes for the three parasite targets and combined with 20X human GAPDH master mix (Applied Biosystems), Taqman multiplex master mix (Applied Biosystems), and cDNA to a final reaction volume of 10 μl. Field samples identified a positive for *P. falciparum* by *varATS* qPCR, no reverse transcriptase controls, amplification controls, and 10-fold parasite RNA dilution standards were assayed in triplicate in 384-well MicroAMP Optical PCR plates (Applied Biosystems). The targets *pfs25, pfs230p, piesp2*, and human *GAPDH* were run in a QuantStudio6 Flex qPCR system (Applied Biosystems) with NFQ-MGB Quencher and VIC, FAM, ABY, and JUN reporter dyes, respectively (**Table S1**). Mustang Purple was selected as the reference dye. Multiplex assay was run under standard cycling conditions: initial denaturation at 95.0°C for 20s (hold stage) followed by 40 cycles of 95.0° C for 1s and 60.0°C for 20s (PCR stage).

### Statistical analysis

All statistical analysis was performed using R version 4.0.1(https://www.r-project.org). Multiple logistic regression was performed with PCR-confirmed gametocytemia as the dependent variable and gender, age (in years), recent bednet use, recent travel, log10 transformed parasite density, and community as independent variables. Plots were rendered using the ggplot2 package. Statistical tests used to determine significance are indicated in tables and figure legends, and p values <0.05 were considered significant.

## RESULTS

A total of 1,354 participants were enrolled across all communities for this study (**Table 1**). The RDTs used for point-of-care diagnosis of asymptomatic infections demonstrated a 4.7% false positive rate using *varATS* qPCR as the reference standard. In contrast, microscopy showed no false positives. Given this, RDT data was not used for subsequent analyses. The prevalence of asymptomatic infections was greatest in Ajigo, followed by Webuye, regardless of diagnostic modality (**Table 1**). Only two asymptomatic infections were detected by PCR in Kapsisywa, and no infections were detected in Kipsamoite. Parasite densities were not statistically different across the four communities (**Table 1**) and did appear to vary with infection prevalence (**Figure S1**). Given the similarities in prevalence of asymptomatic infections in the two highlands communities Kapsisywa and Kipsamoite, they were treated as a single community “Kap-Kip” for all subsequent analyses.

**Table 1.**
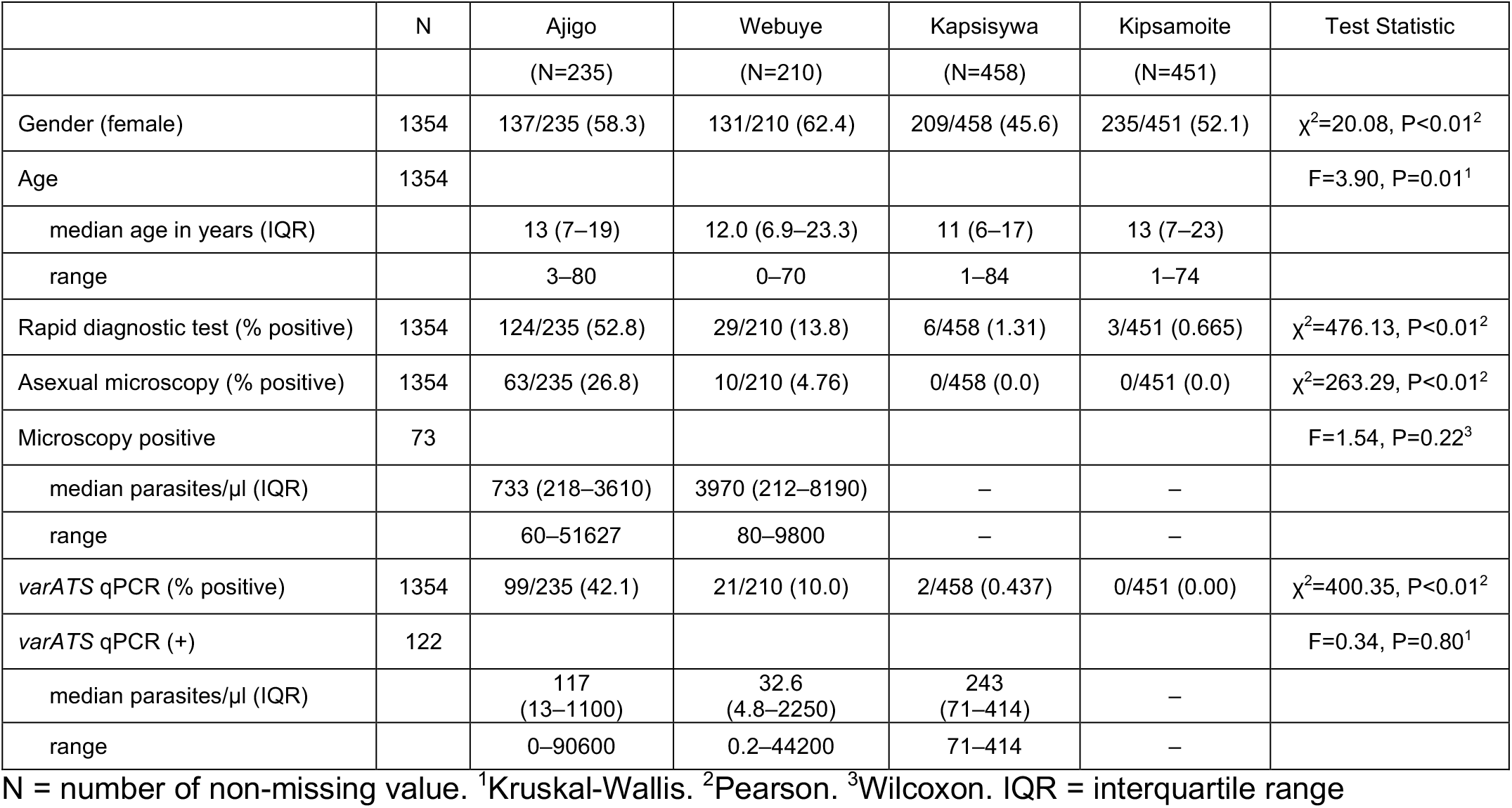
Participant characteristics.

Across all communities, children aged 11-15 years account for the greatest proportion total and sub-microscopic asymptomatic infections (**Figure 1A**). In contrast to the younger age groups, in which the majority of infections are detectable by microscopy, 68% of asymptomatically infected adults >21 years of age have sub-microscopic parasitemia (**Figure 1A**), which suggests acquisition of blood-stage immunity [29, 30]. Similar findings were observed in Ajigo and Webuye when asymptomatic infection prevalence was separated by community with the notable observation that in individuals aged 6-20 years the majority of asymptomatic infections in Ajigo were detectable by microscopy, whereas in Webuye, the majority of infections in this age range were sub-microscopic (**Figure 1B**).

**Figure 1.**
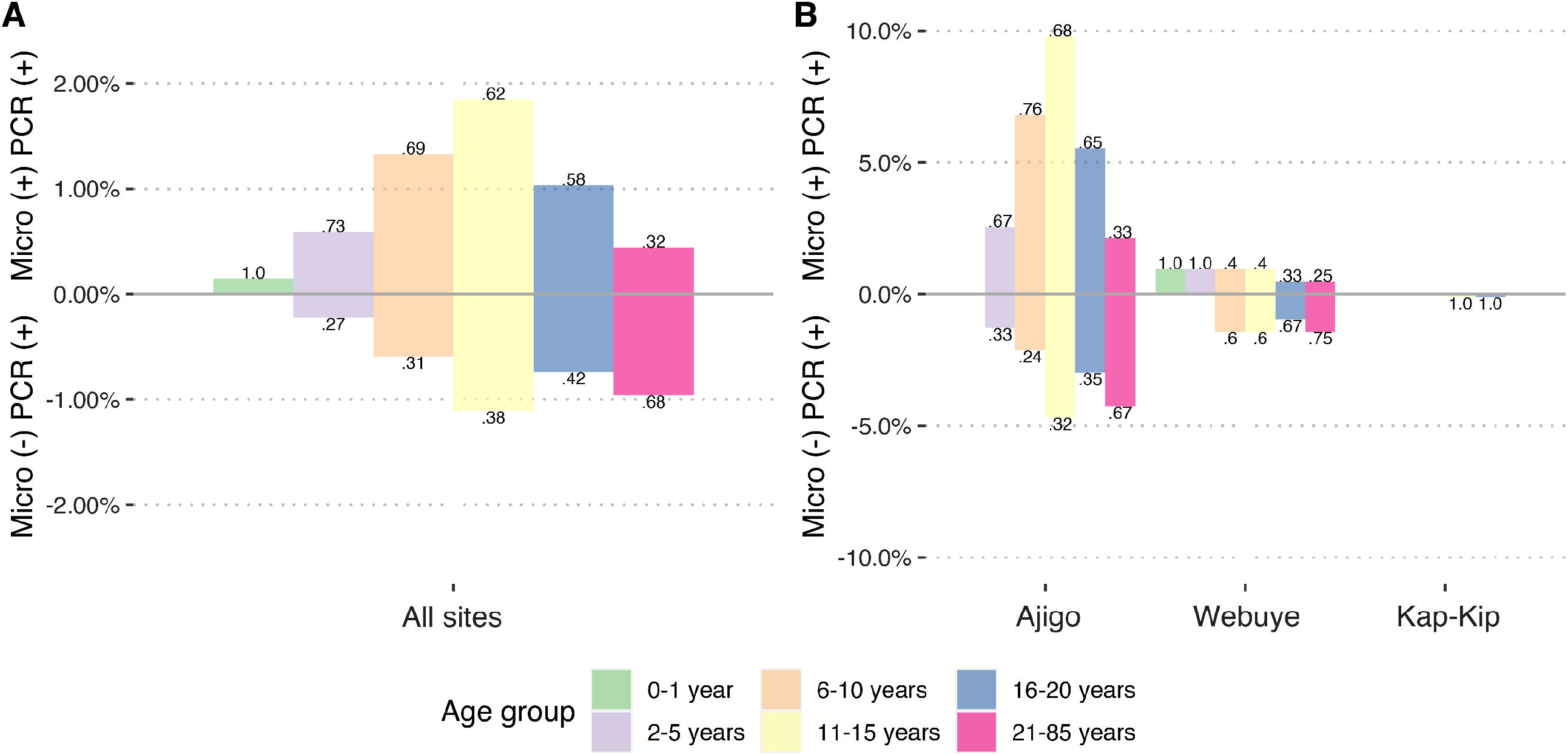
Prevalence of asymptomatic *P. falciparum* infections. Iceberg plot showing microscopy-detectable (above x axis) and sub-microscopic, PCR-detectable (below x axis) *P. falciparum* infections as a proportion of all individuals tested stratified by age group at **(A)** all sites or **(B)** by community. Decimals at above and below each bars represent proportion of microscopy-detectable and sub-microscopic infections in each age group, respectively.

For individuals identified as parasitemic by *varATS* qPCR, we sought to further quantify both asexual parasite densities and sexual parasite densities within the same sample by four-plex RT-qPCR (see Methods). We quantified female and male gametocytes using qRT-PCR targeting *pfs25* and *pfs230p* targets, respectively, and identified 122 of 1354 (9.0%) as having gametocytemia based on the quantifiable expression of either gene. Among individuals with varATS-detectable parasitemia (n=122), there were no significant differences in gender distribution, age, use of ITNs, history of recent travel, or site distribution between those with and without gametocytes by univariate analysis (**Table 2**). To determine the relationship between gametocytemia and total parasitemia, we plotted female and male gametocyte densities estimated by RT-qPCR against estimated asexual parasite density. We initially planned to use parasite densities estimated from *piesp2* Ct values obtained from the same multiplex RT-qPCR assay, which would maintain internal consistency for each sample. However, *piesp2*-derived parasite densities demonstrated poorer correlation with microscopy-determined parasite densities in patent infections and less sensitivity than gDNA-based detection using *varATS* (**Figure S2**). Thus, we used total parasite densities derived from the *varATS*-based assay to approximate asexual parasite densities for the remainder of the study. We included *varATS*-estimated total parasite densities in a multiple logistic regression model that revealed a decreased risk of gametocytemia in the lower transmission communities relative to the high-transmission community of Ajigo and among individuals who reported recent travel (**Table 3**). As expected, increased total parasite densities greatly increased the likelihood of gametocytemia independent of site (**Table 3**).

**Table 2.**
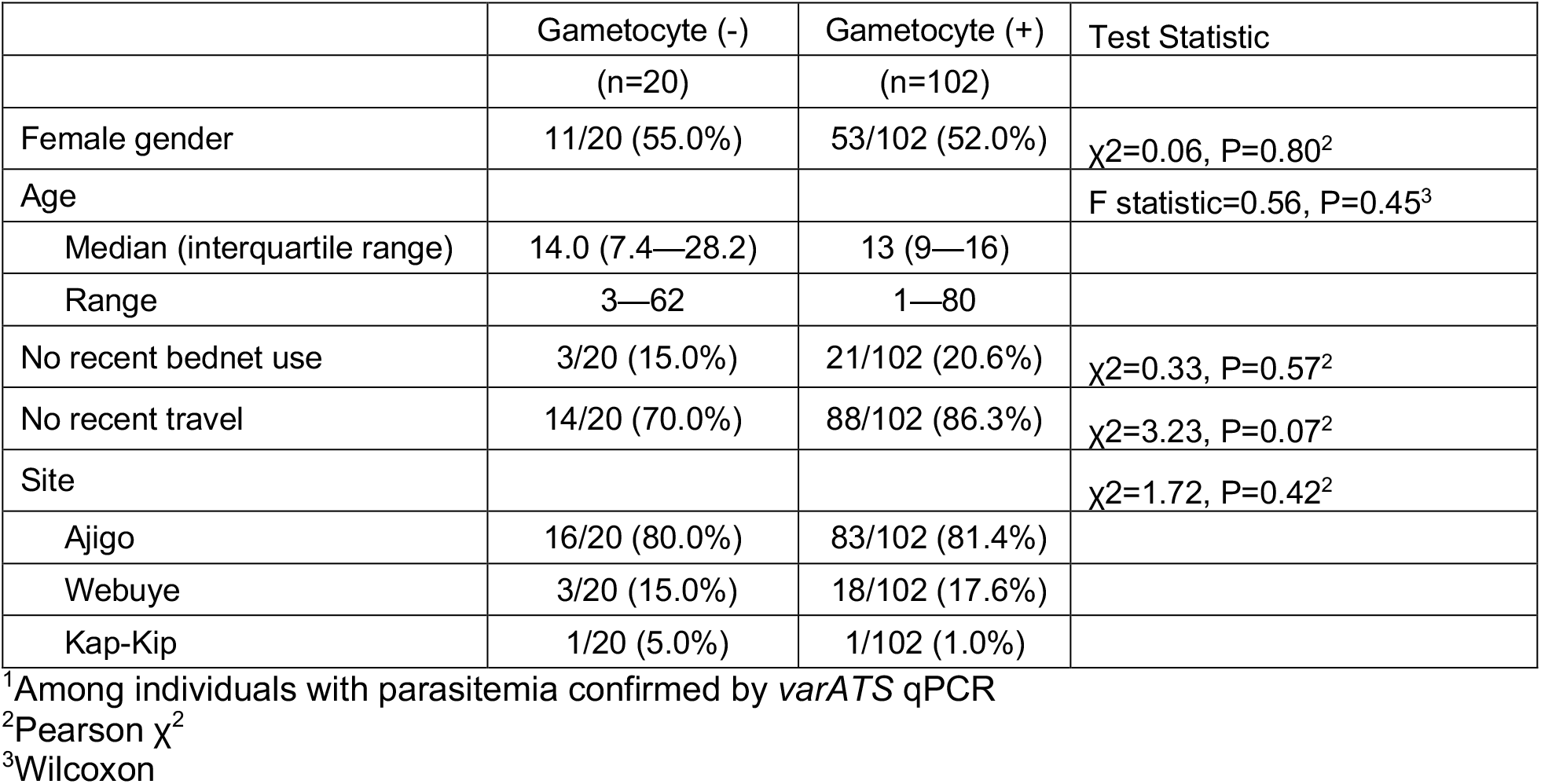
Comparison of gametocyte negative and gametocyte positive individuals.^1^.

**Table 3.** Multiple logistic regression to assess the risk of gametocytemia.

| Variable | OR | 95% CI | p-value | q-value |
| --- | --- | --- | --- | --- |
| Female | 1.00 | 0.55, 1.85 | >0.9 | >0.9 |
| Age | 1.01 | 0.99, 1.03 | 0.2 | 0.3 |
| Recent bednet use (reference: no use) | 0.66 | 0.33, 1.35 | 0.2 | 0.3 |
| Recent travel (reference: no travel) | 0.34 | 0.12, 0.87 | 0.037 | 0.073 |
| $\log_{10}(\text{parasite}/\mu\text{l} + 1)^1$ | 9.40 | 6.11, 15.5 | <0.001 | <0.001 |
| Site (reference: Ajigo) |  |  |  |  |
| Webuye or Kap-Kip | 0.10 | 0.05, 0.18 | <0.001 | <0.001 |

OR = odds ratio; CI = confidence interval
For site, Kap-Kip and Webuye were combined given there were only 2 infected individuals in Kap-Kip.
<sup>1</sup>parasite densities were estimated from *varATS* qPCR Ct values using standard curves (see Methods)

In general, both male and female gametocytemia increased with increasing total parasitemia (**Figure 2A-B**). However, we noted that some individuals with very low total parasite densities had unexpectedly high gametocyte densities. Indeed, individuals with >5 gametocytes/µl were bimodally distributed across a wide range of total parasitemia, which was more marked for *pfs230p* (**Figure 2C-D**). Among individuals with low-density infections (total parasite densities <40 parasites/μl), 28.3% had >5 gametocytes/μl estimated by *pfs230p*, and 20.8% had >5 gametocytes/μl estimated by *pfs25* (**Figure 2E**).

**Figure 2.**
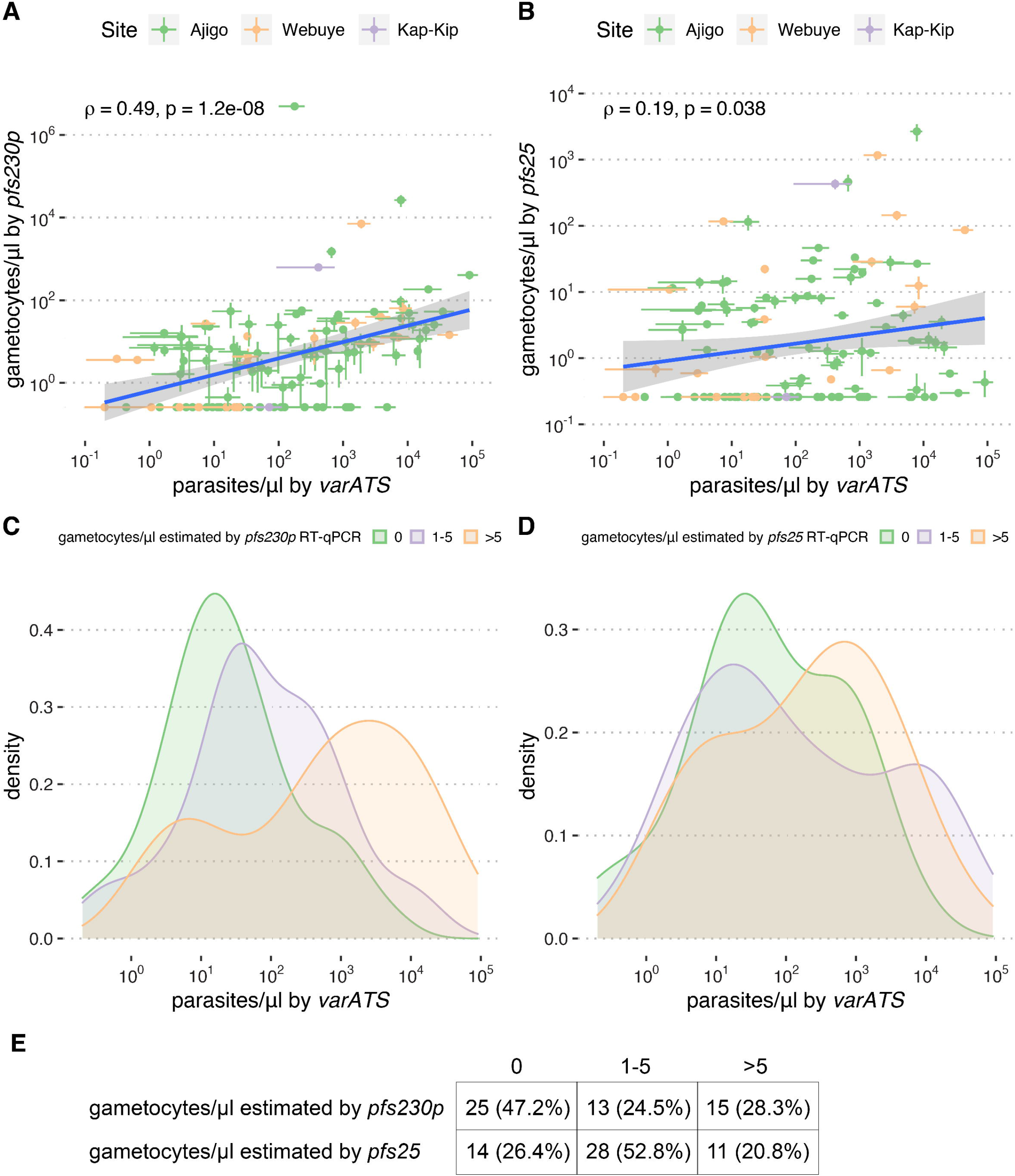
Relationship between total parasitemia and gametocytemia. Spearman’s rank correlations between parasites/μl estimated by *varATS* and gametocytes/μl estimated by (**A**) *pfs230p* or (**B**) *pfs25*. Density plots of parasites/μl by number of gametocytes/μl estimated by (**C**) *pfs230p* or (**D**) *pfs25*. (**E**) Numbers (row proportions) of *varATS*-PCR positive individuals by number of gametocytes/μl.

A substantial proportion (41.7%) of individuals with potential for onward transmission, defined in our study as having at least 1.25 female and four male gametocytes per 2.5 µl of blood (thresholds adapted from a prior study [12] to account for sex-specific gametocytemia overestimation), had qPCR-estimated parasite densities above the detectable limit of conventional, 18s ribosomal RNA-based nested PCR (1 parasite per µl)[21] and below the limit of detection of microscopy (40 parasites per µl), which corresponded well to the actual proportion potential transmitters with submicroscopic infections (36.9%; **Figure 3A-B**). These data provide evidence of a substantial sub-patent infectious reservoir among asymptomatic carriers in these communities but also demonstrates that the vast majority of infections capable of onward transmission can be detectable with conventional molecular diagnostics [12].

**Figure 3.**
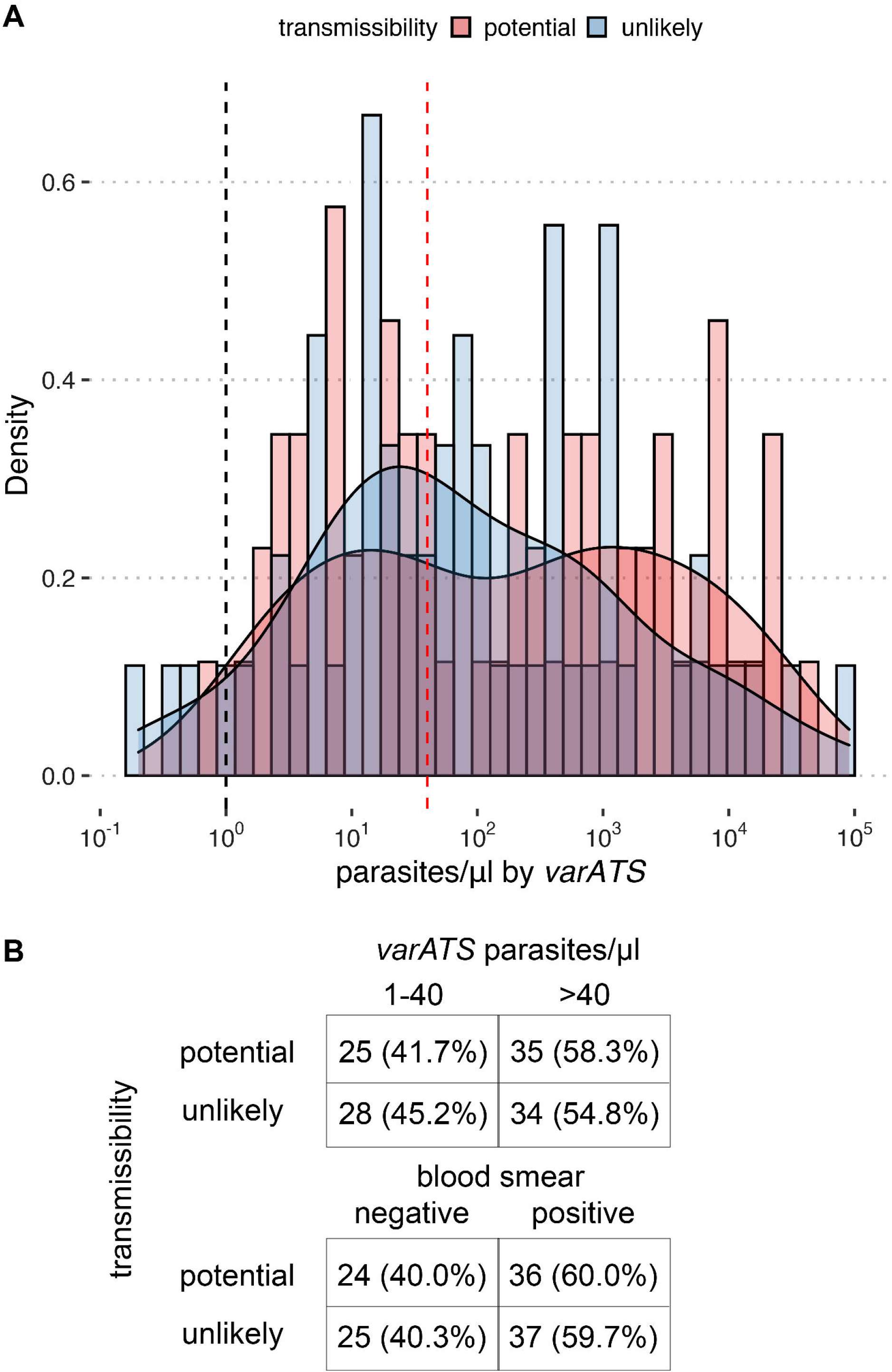
Distribution of individuals with potential for onward transmission. (**A**) Density plots by potential for onward transmission. Limit of detection for conventional molecular diagnostics and microscopy are shown as dashed black and red vertical lines, respectively. (**B**) Numbers (row proportions) of potential and unlikely transmitters by parasite density and blood smear positivity. Individuals with potential for onward transmission are defined in as having at least 1.25 female and four male gametocytes per 2.5 µl of blood, which are increased thresholds adapted from a prior study [12] to account for sex-specific gametocytemia overestimation.

## DISCUSSION

The current descriptive study provides a cross-sectional assessment of asymptomatic *P. falciparum* infections of three communities in western Kenya with differing malaria transmission intensities from August to September 2016. [12]. We confirmed that transmission intensity remained low in Kap-Kip [17, 31], where asymptomatic *P. falciparum* parasitemia was rarely detected by *varATS* qPCR (0.22% prevalence), and established that high malaria transmission occurs in Ajigo, where 42% of individuals had asymptomatic parasitemia. Webuye demonstrated moderate transmission with 10% prevalence of asymptomatic parasitemia, which is lower than what has been previously described at this site [32, 33], possibly reflecting micro-heterogeneity or seasonal differences, as our study was performed during months when rainfall is historically lower in western Kenya.

Our data showing the substantial reservoir among 6-15 year old children in Ajigo and Webuye is consistent with a prior study in the Kakamega district of western Kenya that showed PCR-confirmed asymptomatic *P. falciparum* infections were more prevalent in younger children age 5-14 years (∼34%) relative to older children >14 years (∼9%) [34] but contrasts with a study conducted in a high transmission area (Suba district) that demonstrated the greatest prevalence of blood-smear positive asymptomatic infections in young children <5 years (74% vs. 30-50% in older children) [35]. The differences in relative contribution to the asymptomatic infectious reservoir by age groups may be attributable to intense malaria transmission in Suba, where clinical immunity may be acquired more rapidly, and differences in assay sensitivity.

To determine whether sensitive molecular assays can sufficiently detect the majority of individuals carrying low-density *P. falciparum* infections who are also potentially capable of onward transmission, we assessed the relationship between asexual parasitemia and gametocytemia. We initially intended to correlate gametocyte densities with asexual parasite densities using a multiplex RT-qPCR that would contain targets specific to female gametocytes, male gametocytes, and asexual blood-stage parasites in a single assay, which we had hoped would facilitate comparisons as this strategy eliminates both within subject differences in template preparation and assay variability. However, parasite densities determined using the chosen asexual-specific target *piesp2*, which encodes for parasite-infected erythrocyte surface protein and previously shown to be maximally transcribed in the trophozoite stage in laboratory isolates [24-26, 36], showed weaker correlation with microscopy-determined parasite density than densities derived from *varATS* qPCR using gDNA (**Figure S2**). The weaker correlation for *piesp2* could be due to lower *piesp2* expression in ring stages, which had previously been thought to be the predominant asexual form of *P. falciparum* found in peripheral circulation. However, a recent study in Mali revealed that more developed trophozoite stages were commonly found in asymptomatic *P. falciparum* infections [37]. Expression of *piesp2* could also vary among the field isolates, perhaps due to differential transcriptional regulation related to precise stage at the time of collection or host immune pressure. Although speculative, these potential explanations are intriguing given that antibodies against PIESP2 associate with protection from malaria [38], and PIESP2 has recently been observed to bind to brain microvascular endothelial cells *in vitro* to induce an inflammatory response [39]. Our data, combined with these prior findings, suggest that *piesp2* is a poor target for quantifying asexual parasite densities.

Our observation that a sizable proportion of low-density infections (<40 parasites/μl) had estimated gametocyte densities that would favor onward transmission is consistent with prior studies that demonstrated a considerable sub-microscopic infectious reservoir [4, 10, 40, 41]. Although we only determined the presence of gametocytemia among individuals who were parasitemic by *varATS* qPCR, which had a limit of detection of ∼0.4 parasites/μl using dried blood spots, our data also shows that the proportion of individuals with potential for onward transmission drops off below 10 parasite/μl. This finding is in line with recent studies suggesting that mosquito infectivity occurs primarily when parasitemia is >1 parasite/μl [12, 42], which is the limit of detection of standard molecular diagnostics. Taking together, the main implication is that ultra-sensitive molecular diagnostics capable of detecting infections <1 parasite/μl may not be necessary to achieve significant reductions in malaria transmission using a screen-and-treat strategy.

There are several limitations to our study. The cross-sectional study design provides only a snapshot of infection prevalence in these communities during the relatively dry season and our observations may not be generalizable to the rainy season when malaria transmission is more intense. We estimated gametocyte densities using molecular quantification of male and female gametocyte-specific gene expression as a surrogate of potential for onward transmission and did not directly measure mosquito infectivity using direct or indirect feeding assays. Such a surrogate based solely on gametocyte density along ignores the relative contribution of anti-gametocyte immunity in reducing malaria transmission [43]. Although we use male and female gametocyte targets for gametocyte quantification, we did not differentiate gametocyte sex when determining gametocyte densities by microscopy for our standard curves, which would lead to overestimates of sex-specific gametocytemia. This is especially true for male-specific gametocytemia given that natural infections are biased towards females, with 3-5 times more females [44]. However, we made no assessments using sex ratio in this study. Furthermore, gametocytemia overestimates would affect all samples consistently and thus would not affect our ranked correlation analyses. Importantly, in our determination of number of individuals capable of onward transmission, we adjusted for sex-specific overestimates by using higher thresholds for minimum male and female gametocyte densities.

In summary, our cross-sectional survey of the prevalence and densities of *P. falciparum* infections among asymptomatic individuals in western Kenya provides an assessment of the relationship between parasitemia and gametocytemia in three communities with different transmission intensities. Experimental studies are needed to definitively determine whether the low-density infections in communities such as Ajigo and Webuye contribute significantly to malaria transmission.

## Data Availability

Data will be made to researchers upon request after formal publication.

## ACKNOWLEDGEMENTS

We would like the study participants. We also thank Brian Grimberg (Case Western Reserve University) for assistance with developing the multiplex qPCR assay and Erik Gaskin for review of the literature. This project was funded with support from the Indiana Clinical and Translational Sciences Institute funded, in part by Grant #’s UL1TR001108 and KL2TR001106 from the National Institutes of Health, National Center for Advancing Translational Sciences, Clinical and Translational Sciences Award. The content is solely the responsibility of the authors and does not necessarily represent the official views of the National Institutes of Health.

## AUTHOR CONTRIBUTIONS

GA, WP, CJ, and TT designed the study. GA, EO, AO, and TT coordinated and conducted the field studies. CS, MM, and MG designed and performed the experiments. MM, SN, and TT processed and analyzed the data. CS, WP, CJ, and TT wrote the manuscript. All authors read and approved the final version of the manuscript.

