## Supplementary Material for "The prevalence and density of asymptomatic *Plasmodium falciparum* infections among children and adults in three communities of western Kenya"

Affiliations:
1. Division of Infectious Diseases, Department of Medicine, Indiana University School of Medicine, Indianapolis, Indiana, USA
2. Division of Pediatric Infectious Diseases and Global Health, Department of Pediatrics, Indiana University School of Medicine, Indianapolis, Indiana, USA
3. Center for Global Health Research, Kenya Medical Research Institute, Kisumu, Kenya
4. Jaramogi Oginga Odinga University of Science and Technology, Bondo, Kenya.
5. Reserve Analytics, LLP, Cleveland, Ohio USA
6. School of Medicine, Moi University College of Health Sciences, Eldoret, Kenya
7. Duke Global Health Institute, Duke University, Durham, North Carolina, USA.

### Table S1: Primer and Probe Sequences

| Assay | Primer and Probe name | Sequence | Reference |
|---|---|---|---|
| *varATS* RT qPCR | varATS For | 5' CCCATACACAACCAAYTGGA 3' | |
| | varATS Rev | 5' TTCGCACATATCTCTATGTCTATCT 3' | |
| Multiplex RT qPCR | Pfs230p_2_F | 5' TTGCCTGTTACCTTATTGTCAAG 3' | |
| | Pfs230p_2_R | 5' CCGTCGACATCTTCAGCAT 3' | |
| | Pfs230p_2_IN | 6FAM-CCAGGCTGATGAAGACATCGGG-QSY | |
| | Pfs25 For | 5' GAA ATC CCG TTT CAT ACG CTT G 3' | Wampfler et al. 2013 [24] |
| | qPCR Pfs25 Rev | 5’ AGT TTT AAC AGG ATT GCT TGT ATC TAA 3' | Wampfler et al. 2013 [24] |
| | qPCR Pfs25_probe | VIC-TGT AAG AAT GTA ACT TGT GGT AAC-QSY | adapted from Wampfler et al. 2013 [24] |
| | Pf3D7_ESP_F | 5' TAGGTCAAGCACCTCCAGAA 3' | |
| | Pf3D7_ESP_R | 5' TCATTAGGGTCAAGTGCGTAA 3' | |
| | Pf3D7_ESP_IN | ABY-TGGTTCATTTCCCCAAGCAGGTG-QSY | |

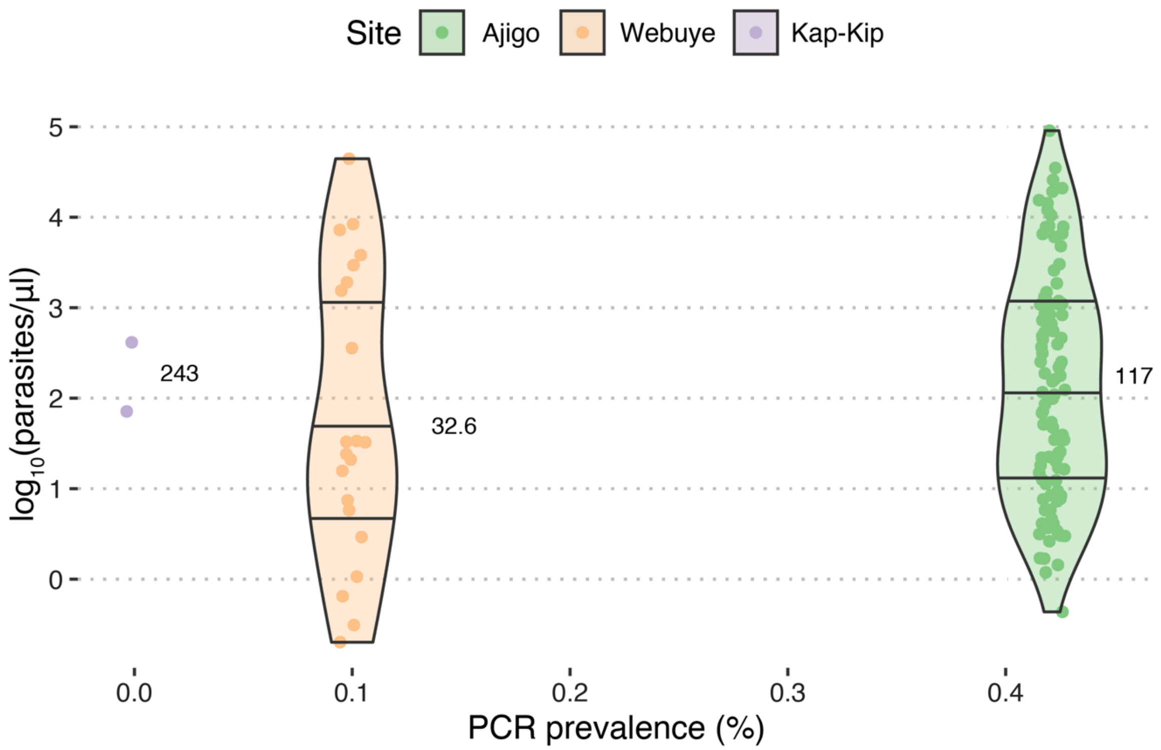


**Figure S1. Relationship between parasite density and prevalence of PCR-detective asymptomatic P. falciparum infections by community.** Median parasite density indicated to right of each violin plot.

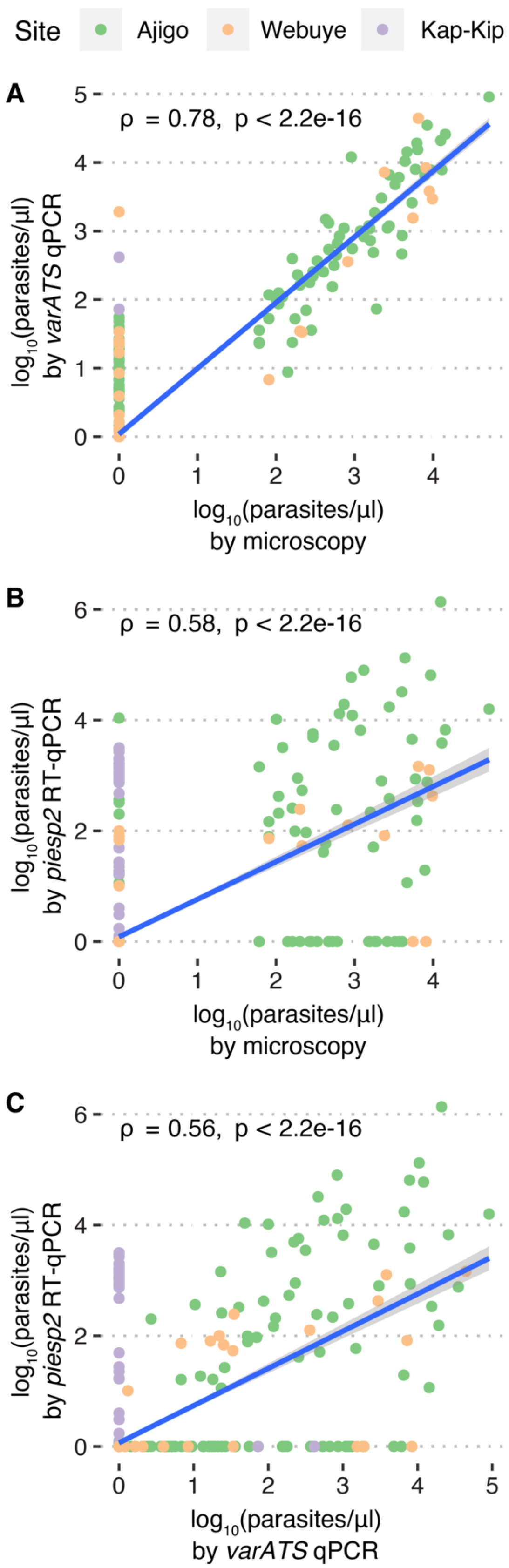


**Figure S2. Correlation between parasite quantification assays.** Spearman's rank correlation coefficients (ρ) were determined between parasite densities estimated by **(A)** *varATS* qPCR and microscopy, **(B)** *piesp2* RT-qPCR and microscopy, and **(C)** *piesp2* RT-qPCR and microscopy. Blue lines are best linear fit lines with 95% confidence intervals.
